# Methods for detection of clusters of observations with an outlying correlation coefficient value

**DOI:** 10.1101/2020.10.12.20211128

**Authors:** Lieven Desmet, David Venet, Laura Trotta, Tomasz Burzykowski, Marc Buyse

## Abstract

Multivariate datasets with a clustered structure are the natural framework for, e.g., multicentre clinical trials. We propose a number of methods aimed at detecting clusters with outlying correlation coefficients. While the methods can be used in a variety of settings, we focus mainly on their application to central statistical monitoring of clinical trials. In particular, we consider the issue of detecting centers (or other clusters of patients such as regions) with outlying correlation coefficients for bivariate data in a multicenter clinical trial. It appears that, in that context, the proposed methods perform well, as we show by using a simulation study and a number of real life datasets.

## 1 Introduction

We focus on the problem of detecting centers with outlying correlation coefficients for bivariate data in a multicenter clinical trial. For example, observing many subjects with a large weight and short height or vice versa in a center may raise suspicion, even if both univariate distributions of height and weight for that center look reasonable. This problem is of interest in the context of central statistical monitoring^1^ (CSM) of clinical trials, where outlying values for the correlation in a center may point to data quality issues that are worth investigating. These issues may include data fabrication, as it is well-known that the multivariate structure of the data is much harder to imitate than the univariate one^2^.

Thus, from a data-fabrication point of view, tests that focus on the multivariate structure of the data are essential in a CSM toolkit. Construction of multivariate tests for central statistical monitoring is challenging as it brings in issues related to dimensionality and parametrization. If many variables are available, the number of dimensions can be daunting. Also, multivariate distributions require more parameters and are harder to test for.

In this paper we focus on a simplified setting: we consider pairs of continuous variables, and assume the bivariate normal distribution as an underlying data-generating mechanism in each center.

The aims of this paper include (*i*) description of suitable data models, both in absence and in presence of outlying centers, (*ii*) formulation of various statistical tests for detection of outlying centers, (*iii*) investigation of the performance of these tests in a simulation study, and, finally, (*iv*) illustration of the methods in a number of real data cases.

The paper is structured as follows. Section 2 describes suitable statistical models and formulates the detection problem. In Section 3, we describe the developed detection methods. Section 4 presents simulation results. In Section 5, we apply the developed methods in three different settings: raw datasets as encountered in central statistical monitoring practice, a biometric multicenter dataset on sports teams, and a set of deliberately fabricated datasets. Section 6 closes the paper with a discussion of different aspects of the described methods, mainly in terms of their performance in different applications.

## 2 Data models and detection problem

### 2.1 Multicenter correlated data

The starting point for formalizing the problem is the description of the null-hypothesis case, i.e., the neutral setting where the centers have a consistent and plausible correlation structure described by a particular statistical model.

We propose a hierarchical model that describes the data in each center and, additionally, the way the correlation structure behaves across centers. In particular, we consider *N* centers. Each center *c* has *n*_*c*_ pairs (*X*_*i*_, *Y*_*i*_), generated from a bivariate normal distribution,

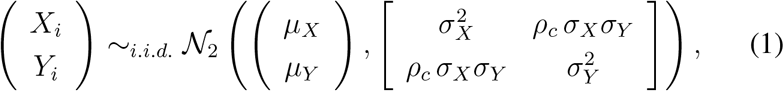

while the center-specific correlation coefficient *ρ*_*c*_ obeys

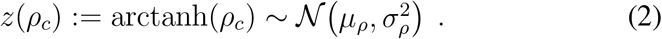

The motivation for this model is two-fold. A bivariate normal distribution is the most general and simplest model that can be assumed to hold, at least approximately, or after transformation, for pairs of continuous random variables. On the other hand, the normal model for the Fisher *z*-transformed correlation parameters is inspired by Fisher’s result on the variability of the Pearson correlation coefficient^3,4^,

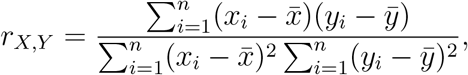

observed across independent replications of a bivariate normal model of size *n* and correlation parameter *ρ*. In particular, *z*(*r*) has approximately a normal distribution with mean *µ*_*ρ*_ = arctanh(*ρ*) and standard deviation 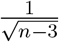. It is therefore natural to allow a normal variability not for *ρ*_*c*_, but for *z*(*ρ*_*c*_).

While equation (1) completes the multicenter structure, it is equation (2) that is crucial in defining the correlation structure across centers, and with respect to which we can assess outlyingness. In equation (2) we allow random deviations from the overall *µ*_*ρ*_ to capture, e.g., unobserved differences in center-specific populations^5,6^.

For wider applicability, we can also consider a more flexible version of equation (1), in which each center *c* has center-specific parameters *µ*_*X,c*_, *µ*_*Y,c*_, *σ*_*X,c*_ and *σ*_*Y,c*_.

However, since the correlations are defined independently for each center, those other center-specific parameters have no influence on the distribution of the center-specific correlations in equation (2).

### 2.2 Detection problem

Our goal is to detect centers that are not compatible with the null model defined by (2). To illustrate the ideas, and for the sake of simplicity, we consider only a specific type of deviation, the so-called *hybrid* model. Of course, the methods proposed are not limited to this specific deviation model.

In the hybrid model we consider a mixture of two types of centers: a majority *N*_0_ that represents normal centers, and a minority *N*_1_ of “contaminant” centers that represent outliers (*N* = *N*_0_ + *N*_1_, *N*_0_ *> N*_1_). All centers follow (2), but with different location parameters: for the majority we set *µ*_*ρ*_ = arctanh(*ρ*_0_), while for the contaminant centers we assume a different location parameter defined by *µ*_*ρ*_ = arctanh(*ρ*_1_).

An example of model densities for *ρ*_*c*_ is shown in Figure 1.

**Figure 1.**
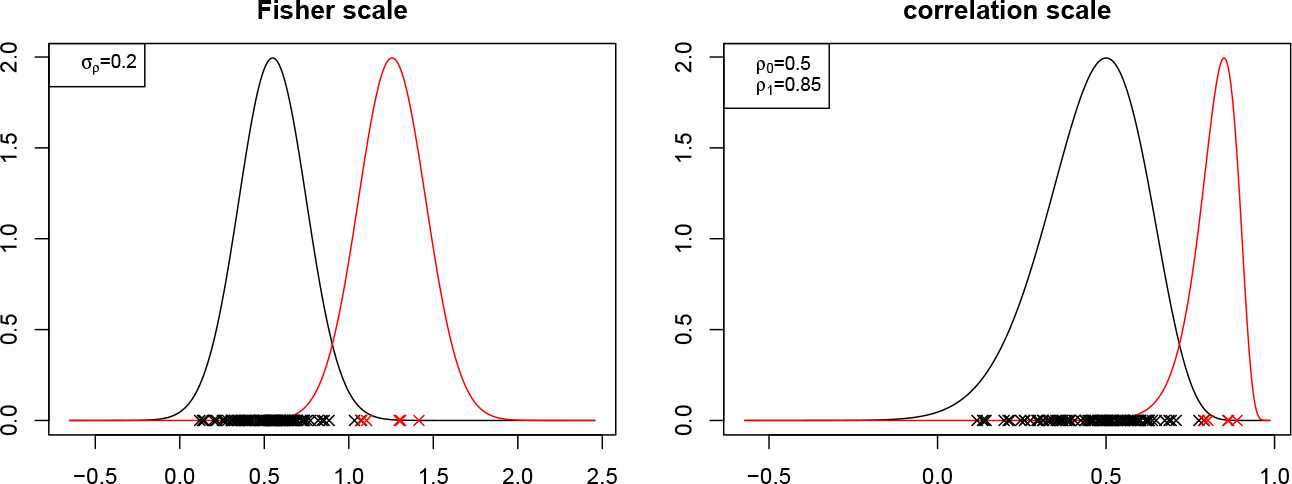
Hybrid setup example. Left panel: normal densities valid for the null model (black solid line, *ρ*_0_ = 0.5 and *σ*_*ρ*_ = 0.2) and alternative model (red solid line, *ρ*_1_ = 0.85 and *σ*_*ρ*_ = 0.2) on the Fisher scale, crosses represent sampled center correlation coefficients (contamination rate is 5%); Right panel: same, but back-transformed to the correlation scale.

The contamination rate is *ϕ* = *N*_1_/(*N*_0_ + *N*_1_). In this setting, the purpose of a correlation test is to detect the *N*_1_ outlying centers among all *N*, by assigning to each center a *p*-value that reflects its compatibility with the null model, and flag a center (i.e., declare it as outlying) when this *p*-value is smaller than the 0.05 threshold (other choices of the significance level are of course possible).

If all centers conform with the null model, the *p*-values should have a uniform distribution and flagging a center that has *p <* 0.05 should lead to a procedure where at most 5% of the centers would be falsely rejected (false positives).

On the other hand, if a few centers conform with an alternative parameter, these should be associated with small *p*-values.

## 3 Detection methods

Broadly speaking, we consider two types of detection methodologies: methods that are directly related to estimation of the null model, which we refer to as Fisher scale methods, and the so-called fixed margin methods with a somewhat different rationale, as described in Section 3.2.

### 3.1 Fisher scale method

This method is based on direct estimation of the model defined by equation (2) on the Fisher scale. In this method we explicitly account for center sizes, and we denote *w*_*c*_ = *n*_*c*_ − 3 and *r*_*c*_ for the correlation coefficient of center *c*.

We obtain estimates 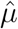 and 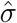 as the arguments that maximize the likelihood

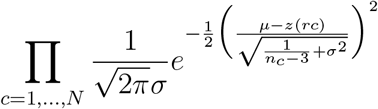

obtained by combining the normal model (2) and the normal approximation to the distribution of Fisher’s *z* transform.

Each center then gets its associated score, given by 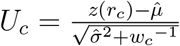, and an associated *p*-value by using the standard normal reference and proceeding as in a two-sided hypothesis test, i.e., we compute *p* = 2 min(*P* (*Z < U*_*c*_), 1 − *P* (*Z > U*_*c*_)), where *Z* is a standard normal variable. As mentioned in Section 2.2, the decision to flag a center is based on checking whether *p <* 0.05.

The general idea of the Fisher scale method is thus to estimate the parameters of model (2) based on the data. It is a reasonable approach conditional on the null model being valid for the data at hand.

For an alternative setting, such as the hybrid model, it is reasonable to make the working assumption that the estimated model is a valid approximation of the null model component provided that the contamination rate is small (say, up to 5 − 10%). Hence, the estimated model can be used as a reference to assess outlyingness.

### 3.2 Fixed margin approach

It is known that in the case of a bivariate normal, the distribution of the sample variances and the distribution of the Pearson correlation coefficient are not independent^7 8^, so that a test on sample variances and a test on correlations would not be independent. This is further compounded by the observation that in real situations, individual centers often include patients from a subpopulation that has different characteristics than the overall population across centers^5^, leading to large differences in variance across centers.

For instance, considering patient height and weight, some centers could be located in cities that have a relatively homogeneous population (hence, little variability in height and weight, and a modest correlation between them), while other centers could have a more heterogeneous population (and a stronger correlation between height and weight). Since the test on correlation is meant to be used as one of many tests, among which tests on marginal variances, it may be warranted to create a test on correlation that is independent on the marginal variance.

The fixed margin approach uncouples the correlation from one of the marginal variances by considering one of the margins (in our example, height or weight) as fixed in the bivariate normal model, thus conditioning the model on the distribution of one of the two characteristics of the center subpopulation.

Specifically, model (1) gives rise to a regression model by conditioning on either of the marginals. For example, by conditioning on *X*, we get the following model

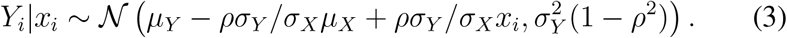

Hogben^8^ describes the variability of the sample correlation coefficient of *x* and *Y* (denoted *r*_*xY*_) in the simple linear regression model: *Y*_*i*_ = *α* + *βx*_*i*_ + *ε*_*i*_, *i* = 1, …*n* with *i.i.d*. normal errors with mean 0 and variance *σ*^2^. In that simple setting, the random quantity 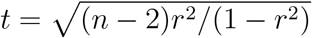 is distributed as a noncentral *t* variable with noncentrality parameter 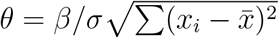 and (*n* − 2) degrees of freedom. Note that the random variability applies to *Y*_*i*_, while the *x*_*i*_’s, and, consequently *θ* are fixed.

Applying this result to our setting implies that 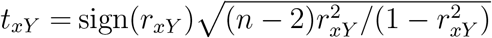 is distributed according to a noncentral *t*-distribution with noncentrality parameter 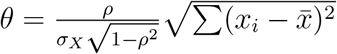 and (*n* − 2) degrees of freedom.

In the practical implementation of the fixed margin approach we introduce a random effect, i.e., a normal model for the center specific *ρ*_*c*_ and proceed as follows. Parameters 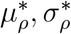 are obtained by numerical optimisation, i.e., as 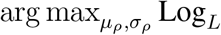, where

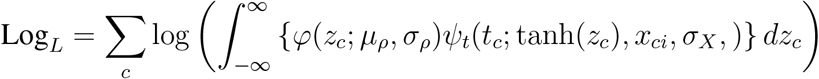

with 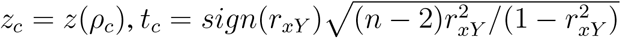 for center *c, φ* the normal density, and *ψ*_*t*_ denoting the abovementioned non-central *t*-distribution. A (two-sided) *p*-value per center is obtained by numerical integration.

The fixed margin approach, by construction, comes in two variants, depending on which of the variables plays the role of the independent (fixed) variable. If we denote the associated *p*-values *p*_*xY*_ (fixing *X*) and *p*_*yX*_ (fixing *Y*), respectively, we can introduce two variants, based on taking the minimum or the maximum of these *p*-values: *p*_max_ = max(*p*_*xY*_, *p*_*yX*_) and *p*_min_ = min(*p*_*xY*_, *p*_*yX*_). Obviously, the former will be the more conservative and the latter the more liberal choice.

## 4 Simulation study

### 4.1 Simulation setup

Simulations are carried out according to the hybrid model introduced in Section 2. We generate data for *N* = *N*_0_ + *N*_1_ centers, where *N*_0_ centers follow model (2) with parameter *ρ*_0_ (null correlation), while *N*_1_ centers follow the same model with a different parameter *ρ*_1_. The contamination rate is *ϕ* = *N*_1_/(*N*_0_ + *N*_1_).

Data are generated according to model (1) with *µ*_*X*_ = *µ*_*Y*_ = 0, *σ*_*X*_ = *σ*_*Y*_ = 1. The number of centers is kept fixed at *N* = 100. Simulation scenarios are constructed by combining different values for the parameters of interest: *ρ*_0_, *ρ*_1_, *σ*_*ρ*_, and *ϕ*. For the sake of generality, and to allow analysis of potential size effects, center sizes are unbalanced. Indeed, in each data generation step, all sizes *n*_*c*_ are drawn at random from a discrete size distribution corresponding to sizes observed in one of three real clinical datasets (Figure 2, in these distributions the sizes smaller than 5 have been replaced by 5). Thus, parameters are chosen as in Table 1.

**Figure 2.**
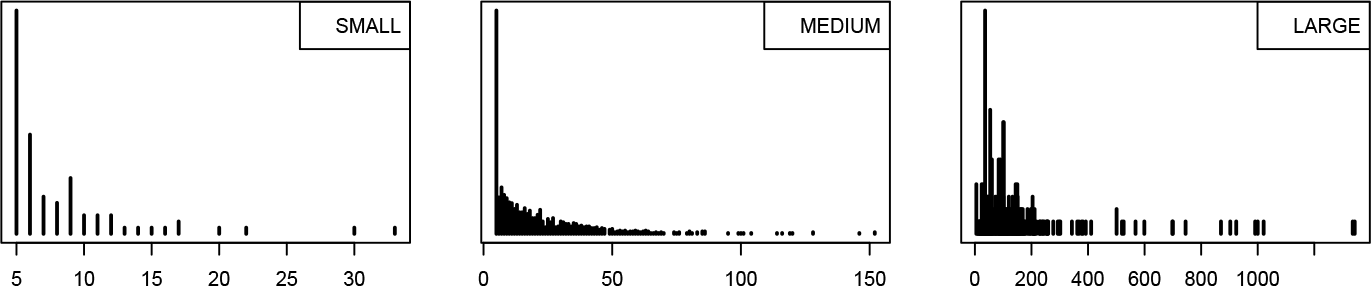
Size distributions: Small, Medium and Large.

**Table 1.**
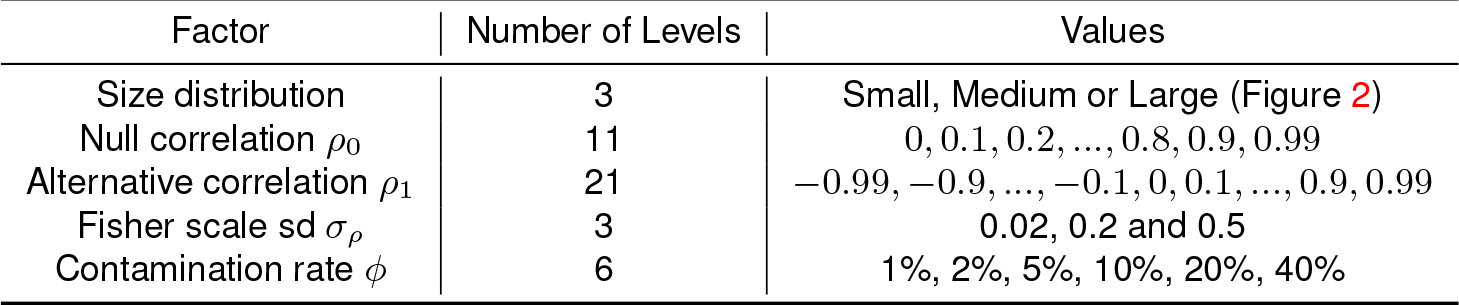
Parameter settings for the simulation study.

| Factor | Number of Levels | Values |
| --- | --- | --- |
| Size distribution | 3 | Small, Medium or Large (Figure 2) |
| Null correlation $\rho_0$ | 11 | 0, 0.1, 0.2, ..., 0.8, 0.9, 0.99 |
| Alternative correlation $\rho_1$ | 21 | -0.99, -0.9, ..., -0.1, 0, 0.1, ..., 0.9, 0.99 |
| Fisher scale sd $\sigma_\rho$ | 3 | 0.02, 0.2 and 0.5 |
| Contamination rate $\phi$ | 6 | 1%, 2%, 5%, 10%, 20%, 40% |

### 4.2 Method evaluation

All methods assign a *p*-value to each center. The center is flagged as an outlying one when its assigned *p*-value is smaller than the 0.05 threshold. The distribution of *p*-values should be reasonably uniform under the null model.

Performance is measured in terms of power and specificity across a number *n*_*sim*_ of replications of the cycle: *multicenter data generation - p-value computation (several methods) - binary detection decision per center*.

More precisely, in each replication the number of true positive, true negative, false positive, and false negative center detections are computed for the multicenter table at hand and these numbers are summed across replications giving rise to totals *TP, TN, FP*, and *FN* respectively. Then power is then computed as *TP* /(*TP* + *FN*) and the specificity as *TN* /(*FP* + *TN*).

The precision of the power estimate can be assessed by using a binomial model: an upper bound for the standard deviation of the power estimate is provided by the expression 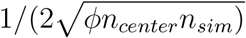. For example, with *n*_*center*_ = 100, contamination rate *ϕ* = 0.02, and *n*_*sim*_ = 100, the bound is 0.035 (standard deviation scale).

### 4.3 Selected results and discussion

When assessing the performance and validity of different detection procedures, we examine a number of properties: (*i*) power should be positively correlated with the absolute difference between *ρ*_1_ and *ρ*_0_ (bearing in mind that, in addition to the individual values *ρ*_0_ and *ρ*_1_, the random effect standard deviation *σ*_*ρ*_, and center sizes also play a role), and (*ii*) specificity should be close to or above 95% across all scenarios (in line with the 5% threshold). Additionally, (*iii*) power should be high for small contamination rates and decrease with increasing rate (at *ϕ* =40%, say, one might argue that it can no longer be considered contamination). Taking into account the unbalanced nature of center-specific sample sizes in clinical trials, in addition, (*iv*) the correlation test should not be biased by center size, i.e., the probability for a center to be detected as outlying should not be influenced by its size.

Figures 3 and 4 present performance curves for the Fisher scale and fixed margin methods as a function of the alternative correlation *ρ*_1_ for a fixed null correlation *ρ*_0_ and given standard deviation and distribution of the center sizes. These figures confirm that properties (*i*) and (*ii*) are met for both tests in these simulations for a small contamination rate, which is the situation of greatest interest in practice.

**Figure 3.**
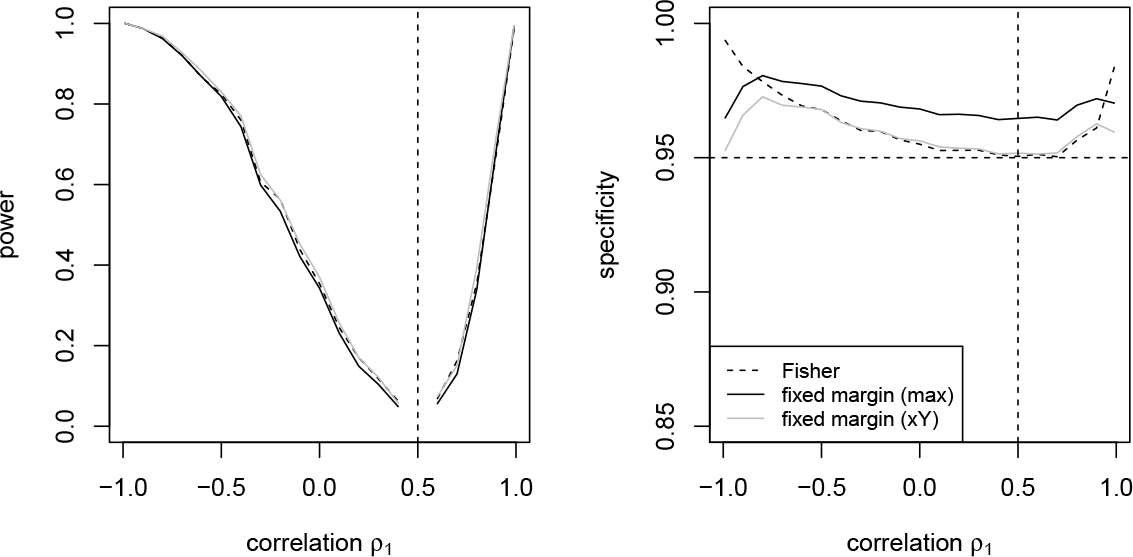
Performance curves: dashed lines for Fisher method (black) and solid lines for fixed margin (black: maximum, grey: fixing X). Moderate variance *σ*_*ρ*_ = 0.2; Medium sizes; 2% contamination; departures w.r.t. *ρ*_0_ = 0.5, *n*_*sim*_ = 400.

**Figure 4.**
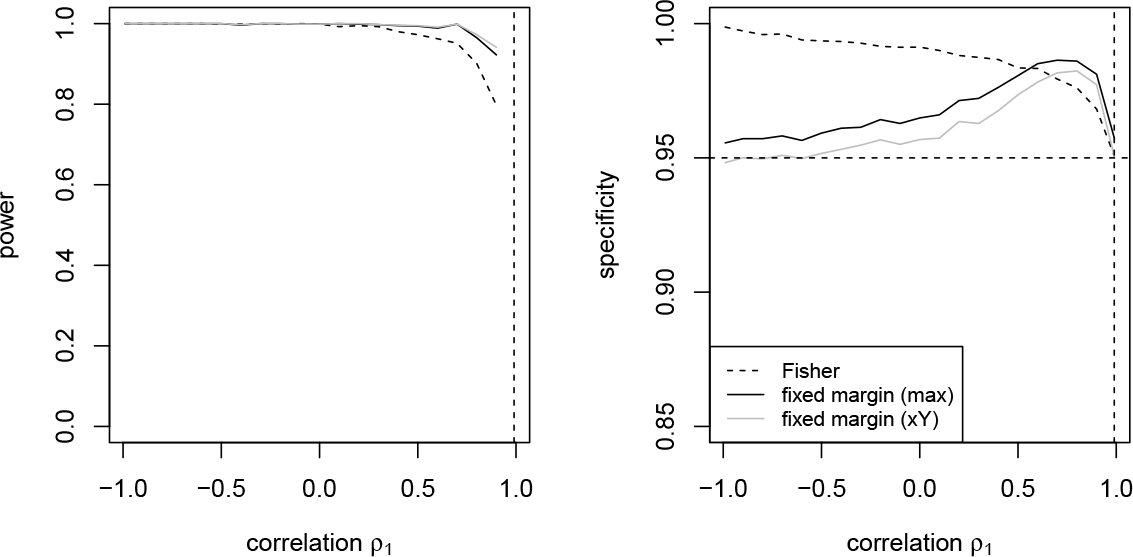
Performance curves: dashed lines for Fisher methods (black) and solid lines for fixed margin (black: maximum, grey: fixing X). Moderate variance *σ*_*ρ*_ = 0.2; Medium sizes; 2% contamination; departures w.r.t. *ρ*_0_ = 0.99, *n*_*sim*_ = 400.

Figures 5, 6 and Supplementary Figure S1 present a summary of the results from a broader set of simulations using multiple scenarios to investigate properties properties (*i*) to (*iii*). These results refer to simulations with *n*_*sim*_ = 100 replications and the grid of correlation values is a subset of that reported in Table 1, namely −0.99, −0.8, −0.5, 0, 0.5, 0.8, 0.99 for *ρ*_1_ and 0, 0.5, 0.8, 0.99 for *ρ*_0_.

**Figure 5.**
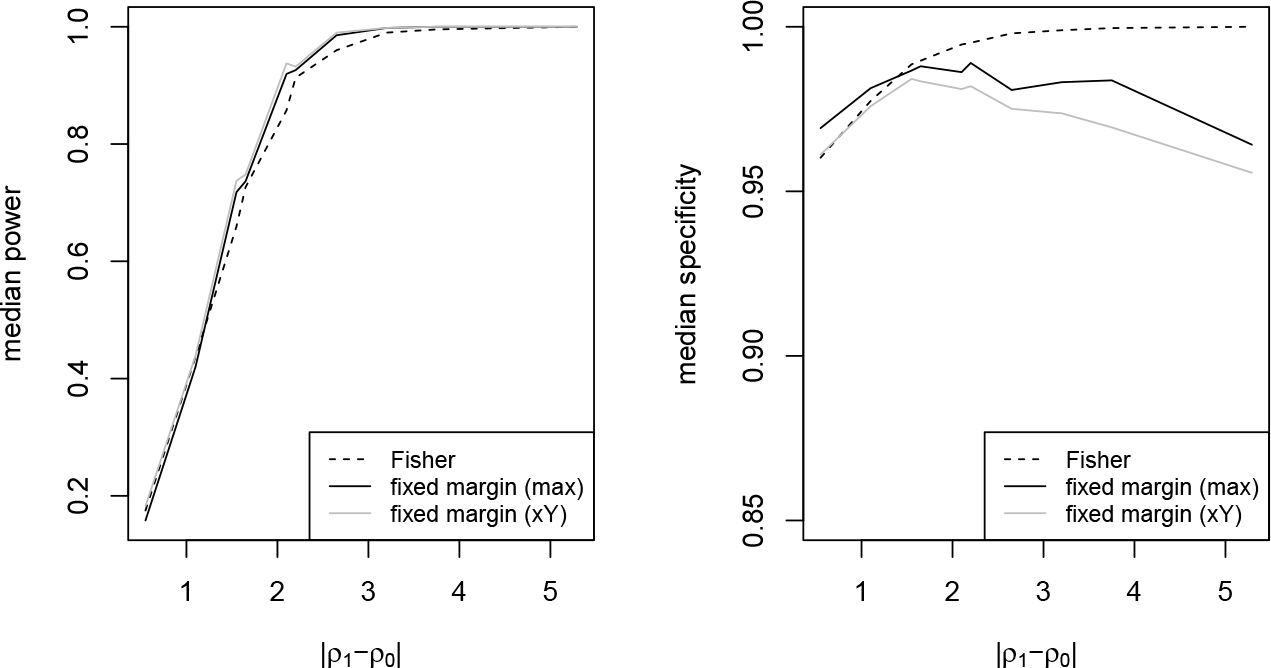
Median performance, aggregated across scenarios for a given difference |*ρ*_0_ − *ρ*_1_|. Power curves start at 20% approximately while specificity curves are above 95% throughout.

**Figure 6.**
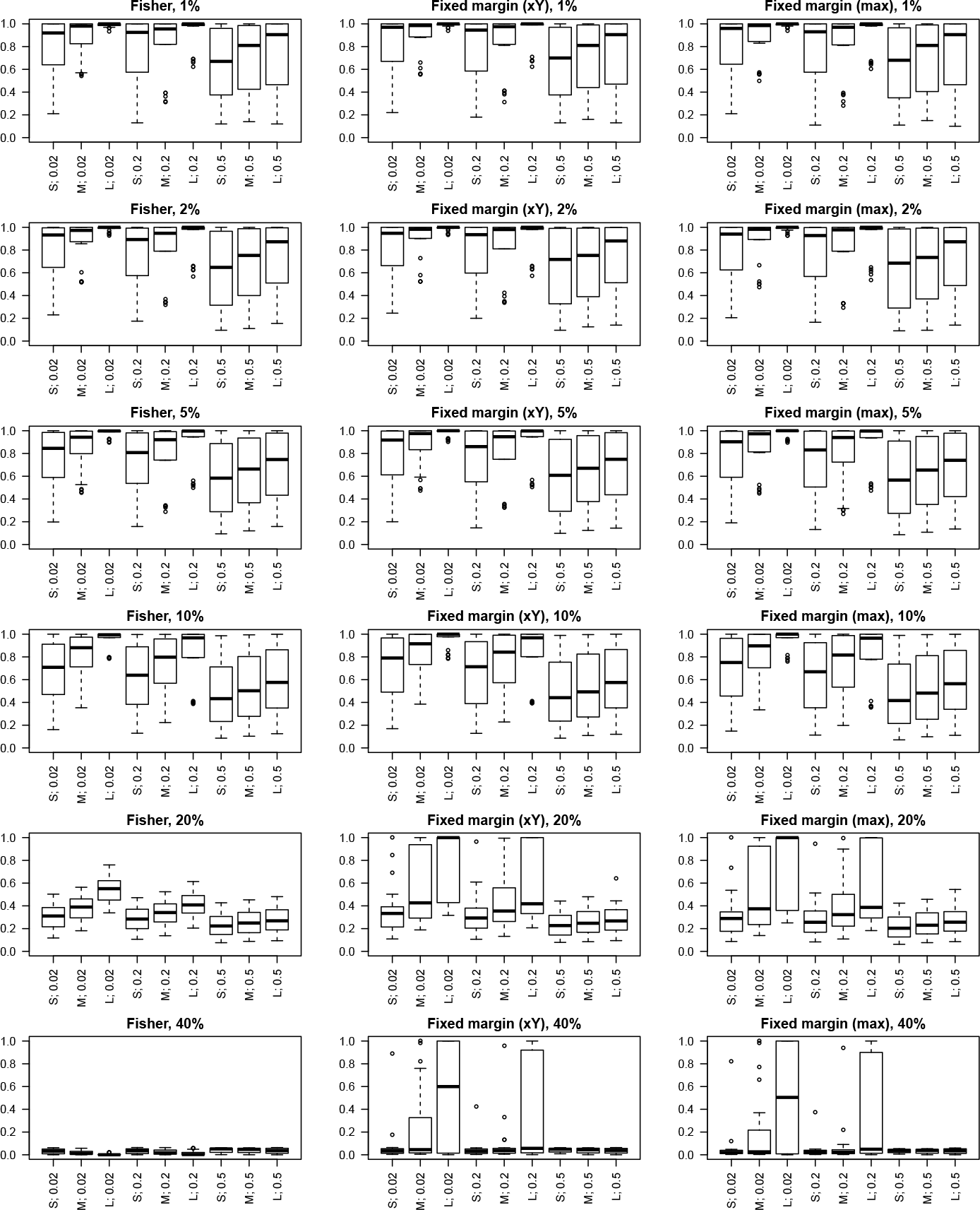
Power boxplots in a matrix for different levels of contamination (rows) for Fisher method (1st column) and fixed margin method (2nd column: fixing X; 3rd column: maximum version). Individual boxplots per panel are identified by X-axis labels: center size indicator (Figure 2) followed by Fisher scale standard deviation.

In Figure 5, the effect of absolute magnitude of the signal, defined as |*ρ*_1_ − *ρ*_0_|, on both power and specificity is represented in terms of the median across different scenarios and different matching (*ρ*_0_, *ρ*_1_) pairs for the same |*ρ*_1_ − *ρ*_0_| value. In spite of being a rough summarisation, the effect of increasing |*ρ*_1_ − *ρ*_0_| is clearly monotonic before the curves level off close to 100%. There is little difference between the reported methods in terms of power and their specificity is conservatively controlled.

Figure 6 (for power) and Supplementary Figure S1 (for specificity) depict the combined effect of contamination rate, sizes distribution, and random effect size (*σ*_*ρ*_) across all available (*ρ*_0_, *ρ*_1_) hypotheses for the Fisher scale method, the fixed margin method (fixing *x*) and its maximum variant. For the power it is useful to make a distinction between two contamination ranges, say 1% to 5% and 10% to 40%.

Focusing on the low contamination range, the sample size effect is quite strong, for a given *σ*_*ρ*_. For a given sample size, increasing *σ*_*ρ*_ introduces more variability in the power estimates and decreases overall power. Remarkably, the reported methods are rather similar and achieve a good level of power at these small contamination rates, with overall performance decreasing slightly with an increasing contamination rate.

In the higher contamination range, power decreases more sharply, but not evenly across methods. For the Fisher scale method it decreases rather uniformly towards 0, whereas for the fixed margin method we see substantial power in some scenarios, even at 40% contamination rate (mostly in cases where either *ρ*_0_ or *ρ*_1_ equals 0.99 or −0.99).

In terms of the specificity all methods are conservative, i.e., their specificity remains consistently above the 95% bound. In Supplementary Figure S1, we can observe increasing specificity with increasing contamination rate.

In order to assess property (*iv*), we compared distributions of the sizes of the detected and the undetected centers. As a consequence of the setup of our experiment, centers with the alternative correlation should have the same size distribution as the original size distribution, shown in Figure 2. The analysis led to the conclusion that all methods comply reasonably well with the property, as shown in Supplementary Figure S2.

In view of its robustness, the test based on *p*_*max*_ turns out to be the best option among the fixed margin test variants, as taking the maximum implies only a negligible loss of power with respect to the tests based on *p*_*xY*_ or *p*_*yX*_.

In conclusion, in the reported simulations, both the Fisher scale and Fixed margin methods exhibit sufficient power to detect small amounts of contamination, while they conservatively control the type I error probability.

## 5 Applications and clinical examples

In this section we apply the proposed tests to actual datasets. The Fisher scale test and the maximum variant of the fixed margin test now become our default tests. We will refer to them as the *Fisher test* and *Fixed margin test*, respectively, and we refer to their *p*-values with *p*_*F*_ and *p*_*H*_ (in honour of Hogben’s result), respectively.

### 5.1 Baseball players example

We apply the methods to a dataset containing height (in cm) and weight (in kg) of 1033 baseball players from 30 Major League teams. The data are publicly available as part of the Statistics Online Computational Resource (SOCR) project of UCLA university^12^.

This dataset has a different size distribution than usually seen in clinical trials, as teams are typically of size around 30 (ranging from 28 to 37) and the setup is thus reasonably balanced without small or large “centers”. From the descriptive statistics (Table 2) we can conclude that the marginal distributions are quite comparable across teams. In particular, standard deviations vary from team to team by no more than a factor 2 and this holds both for height and weight. There is more variability in terms of observed team-specific correlations. Figure 7 contrasts the scatterplots of the aggregated data (overall correlation is 0.5319) and three selected teams with extreme correlations: MLW (0.2147), PIT (0.7198), and WAS (0.7233). In MLW and WAS, the correlation is in fact driven by one extreme observation. Indeed, WAS happens to include the tallest player in Major League Baseball history, while data for the other player (118 kg for 183 cm) is on the edge of the cloud, but still plausible. In PIT, the correlation seems to be driven by a concentration of players in a fairly linear configuration in the center of the cloud.

**Table 2.**
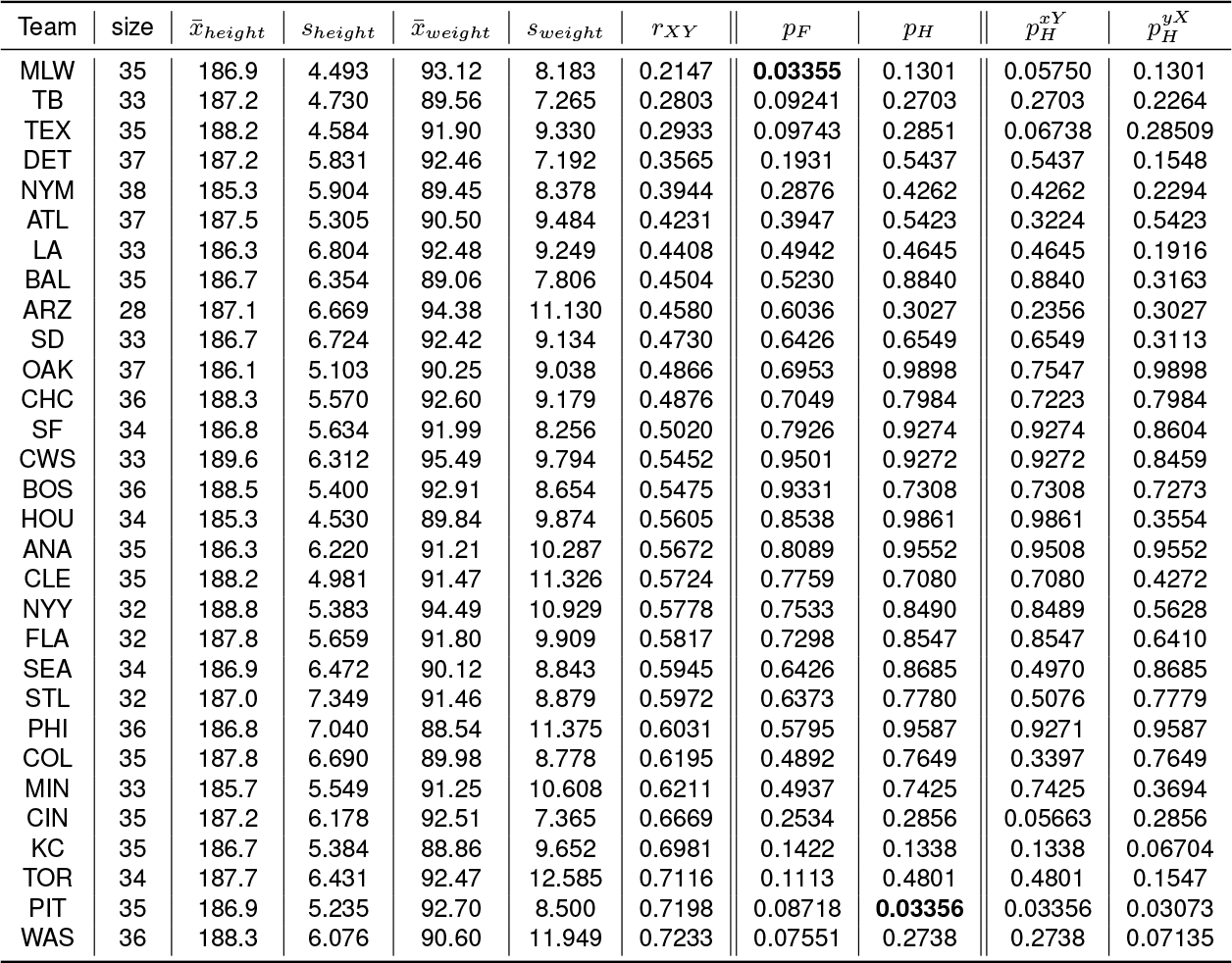
Baseball teams data: means, sample standard deviations and Pearson correlations of the teams and *p*-values for different tests.

**Figure 7.**
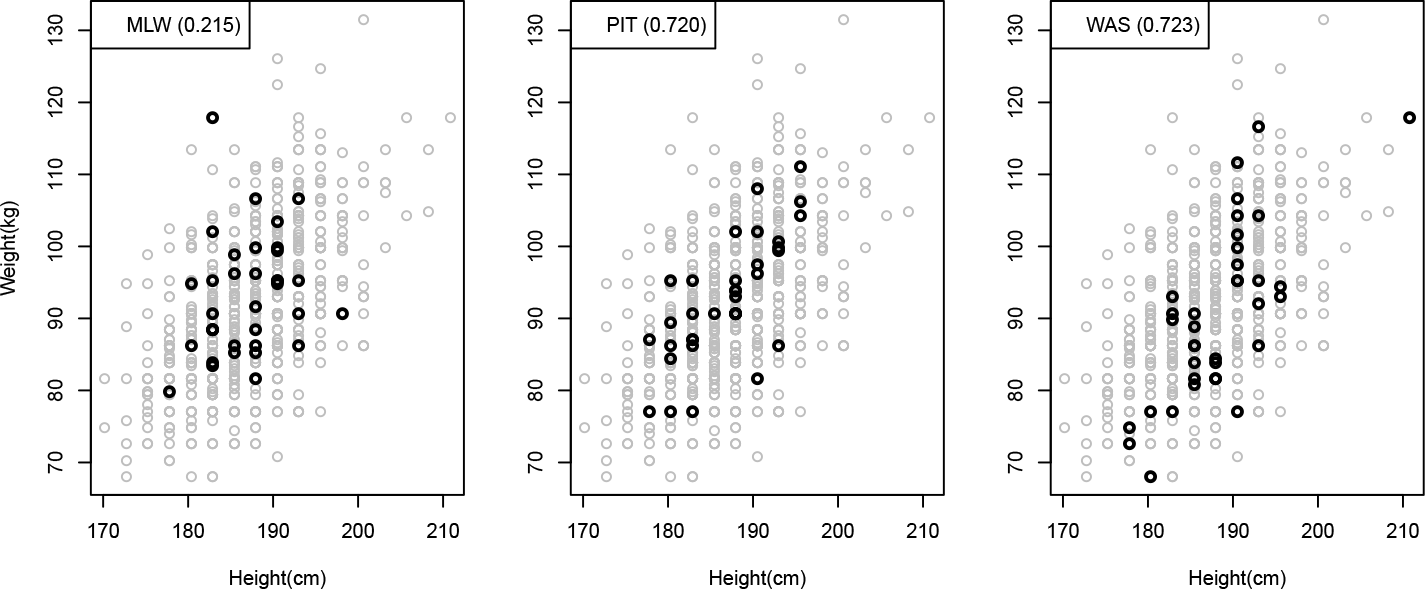
Height-weight data overall (grey dots) and in teams MLW, PIT and WAS (black dots).

The *p*-values for the two proposed tests are shown in Table 2 (which also shows the Fixed margin variants with conditioning on *X* and *Y*, respectively). Both the Fisher and the Fixed margin test each point to one detection (using the 0.05 significance level) among the 30 teams, where the detections concern teams with an extreme correlation already mentioned above. The distribution of *p*-values (not shown here) was reasonably uniform, as expected under the null. Thus, the flagged teams are probably false positives as we have no reason to believe that the data are not genuine or inaccurate, provided of course that the hypothesized data-model is valid for this example. Note that, moreover, reported *p*-values are not corrected for multiple testing.

### 5.2 Detection of fabricated datasets

Akhtar-Danesh and Dehghan-Kooshkghazi^10^ investigated how the correlation structure differs between real and fabricated clinical datasets. They considered two real bivariate datasets, and for each one they had 34 faculty members make up a dataset (with 40 ficticious patients) mimicking the original correlation structure as closely as possible.

It turns out that the investigators, in spite of detailed instructions and knowledge of the original data, failed to reproduce the data structure. Of interest to our research was the question whether these datasets could be identified as fabricated by a correlation test.

We considered the two cases given in Table 3, and designed a numerical experiment. In particular, we constructed hybrid multicenter datasets, in which 19 centers were simulated according to the bivariate normal with parameters as in the table (to be considered as the null model) and one center was taken from the list of fabricated centers (kindly supplied by the authors of ^10^).

**Table 3.** Characteristics of the two original datasets.

| sample (size) | variable X | variable Y | correlation |
| --- | --- | --- | --- |
| Students (65) | Height (in cm, $\bar{x}$=159.5, $s_X$=7.2) | Weight (in kg, $\bar{y}$=54.5, $s_Y$=9.2) | $r$=0.43 |
| Newborn boys (637) | Gestational Age (in weeks, $\bar{x}$=40.1, $s_X$=1.0) | Birth Weight (in g, $\bar{y}$=3277, $s_Y$=443) | $r$=0.031 |

This simulation was repeated 100 times for each of the 34 fabricated datasets. Based on a detection threshold of 0.05, power to detect the fabricated center and specificity were computed per hybrid dataset and reported in Supplementary Figure S3 (averages across 100 replications).

We concluded that the different correlation tests had comparable power. They also detected the majority of the fabricated centers (in half of the scenarios with a power of at least 90%, and in two thirds of them with at least 50% power). The centers with a correlation closest to that of the null model were, as expected, the most difficult to detect.

An investigation of the use of the Fixed margin test, combined with other tests on continuous variables in a central statistical monitoring approach on these data, is the subject of a separate manuscript^11^. Indeed, when data are fabricated in one or a few centers, this may also be visible on the univariate level in terms of discrepancies in the means or standard deviations of the variables *X* or *Y* across centers. While the Fixed margin test would have detected about half of the 34 fabricated datasets, use of all the data features would have detected all of them^11^.

### 5.3 Clinical examples

We analyse a few datasets from existing clinical trials in different therapeutic fields. Clinical datasets typically have a large number of continuous variables, hence a huge number of pairs of variables to be tested, in multiple centers of different sizes. In addition, contrary to simulated data, real clinical data cannot be assumed to strictly follow an underlying bivariate normal model. In particular, there may be some extent of disparity in the center means or scales (i.e. standard deviations of the *X* or *Y* variable).

For ease of interpretation, we will consider two pairs of continuous variables that are known to be positively correlated: diastolic and systolic blood pressure, and the height-weight pair, which we revisit. These variables are available in most clinical trials as part of the vital signs data collected for each patient at each visit.

The main question we try to answer is: which centers are flagged and can this be interpreted in terms of the correlation structure, and ideally in terms of underlying phenomena on the domain level.

Secondly, we also investigate whether the different tests for outlying correlation yield consistent results.

To obtain insight into the correlation structure of this type of data, we can try to rely on reference ranges reported in literature or obtained from publicly available datasets. In spite of compelling biological reasons for the considered pairs of variables to be positively associated, observed correlation values are often lower than one might expect. In the height-weight pair, the observed correlation is directly influenced by the characteristics of the population. Islam et al.^13^ report a correlation of 0.435 in a group of 354 male students and of 0.319 in a group of 285 female students of the same age range (18 to 25). Among (very fit) professional baseball players, a larger correlation was obtained: 0.532 across teams. In the blood pressure pair, both quantities are affected not only by baseline characteristics and underlying conditions of the patients, but also by temporary conditions of stress, level of activity etc. However, observed correlation tends to be higher than in the height-weight pair. Gavish et al.^9^ report a correlation of 0.74 in a study with 140 patients (age 56 +/- 17 years, 45% men).

We use example datasets from real clinical trials with characteristics as given in Table 4 (only centers of size minimum 5 and non-zero variance in both variables are included, after discarding incomplete cases).

**Table 4.** Pairs of variables and characteristics of the trials they are extracted from.

| Trial name | Pair | #Patients | #Centers | Sizes | Median size | mean correlation |
| --- | --- | --- | --- | --- | --- | --- |
| A | diastolic-systolic blood pressure | 4659 | 43 | 32-226 | 100 | 0.5688 |
| B | diastolic-systolic blood pressure | 2711 | 240 | 5-140 | 8 | 0.5646 |
| B | height-weight | 2785 | 245 | 5-146 | 8 | 0.2554 |
| C | diastolic-systolic blood pressure | 632 | 59 | 5-32 | 9 | 0.6759 |

As expected, center size plays a major role. Figure 8 presents the relationship between correlation and center size across centers for all four trials. In trial B, that has many centers, the scatterplot reveals a funnel-like pattern. This reflects the variability present in the Pearson correlation: smaller centers have more variability (wide opening along the Y-axis), larger centers less (defining a narrow tube pointing to the right).

**Figure 8.**
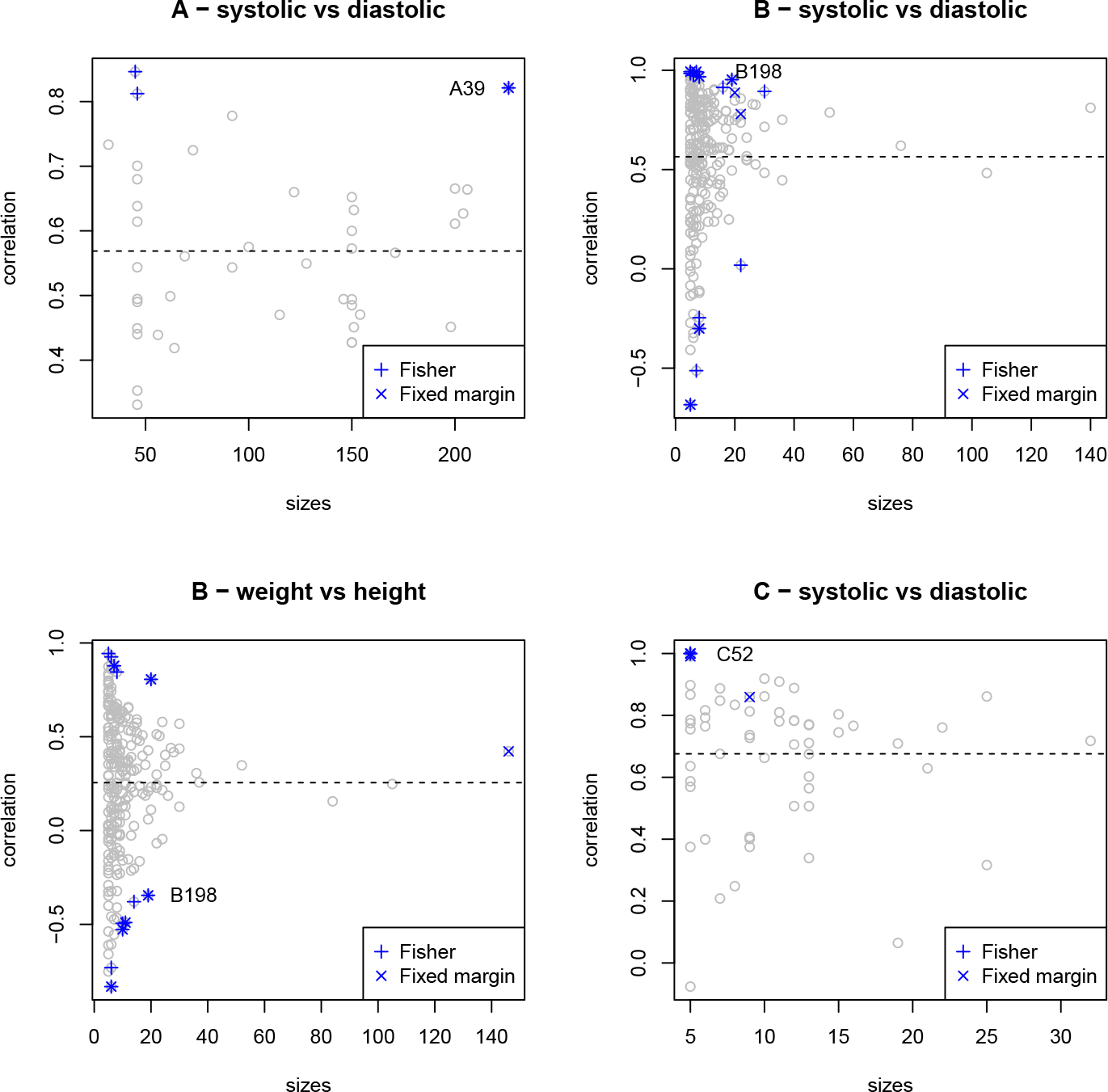
The relationship between center sizes and correlation in the example datasets, flagged centers are marked for the Fisher and the Fixed margin tests.

In trial C with small centers we see more variability on the Y-axis, and in trial A with larger centers we have less variability as seen on the Y-axis (in these two cases only part of the funnel is observed).

It is indeed across the edges of the funnel that centers are detected: centers with a too high or too low correlation, taking into account their size.

Figure 9 presents scatterplots of several particular cases for further detail.

**Figure 9.**
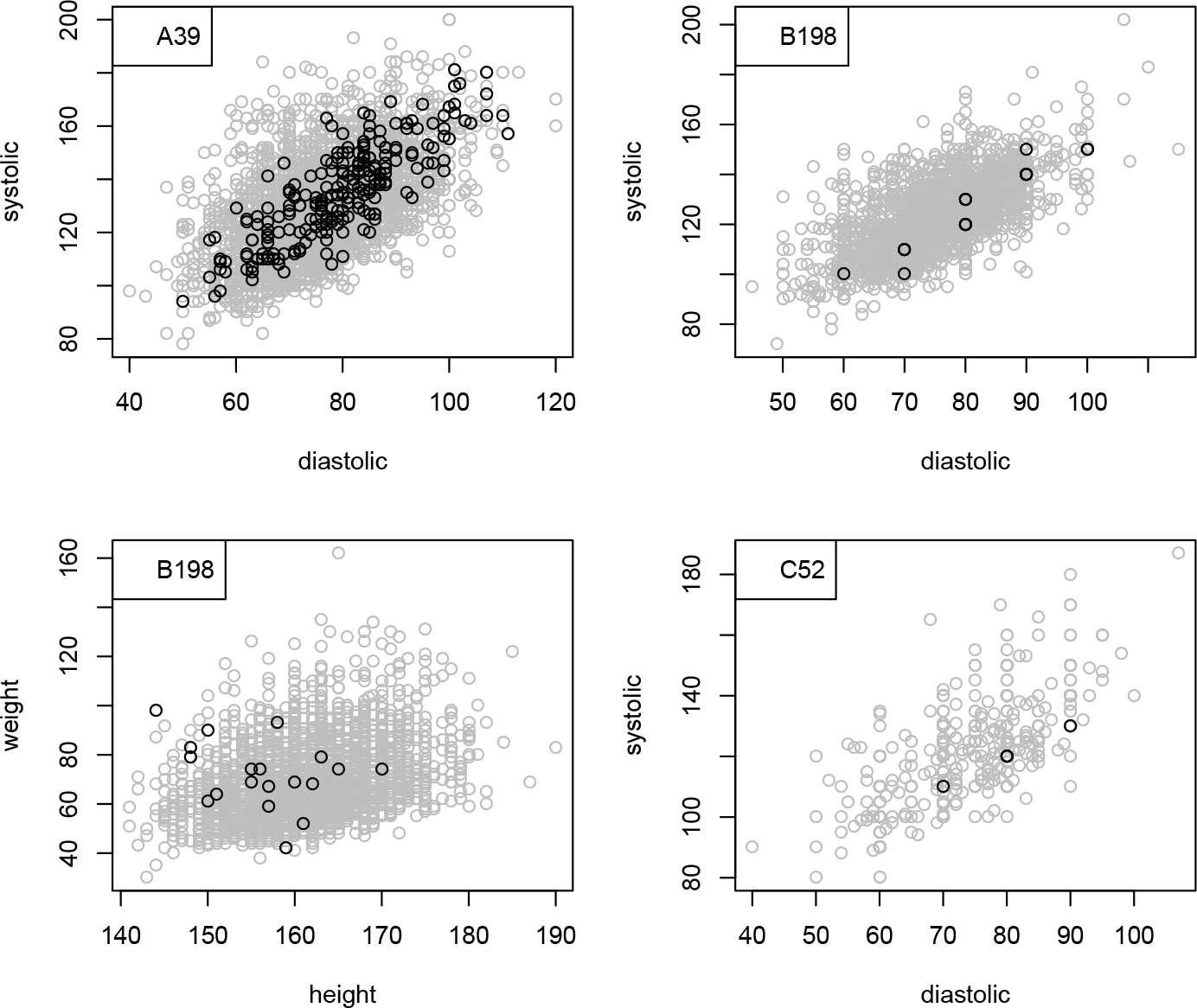
Four typical cases that are detected with both tests. Center data (black) are contrasted with overall cloud (grey).

In trial A, a large center (A39) is detected with both methods with *p*_*H*_ = 0.034 and *p*_*F*_ = 0.0059 (size=226, correlation of 0.8211). In trial B, a center (B198) of size 19 is detected for both pairs of variables. The pattern in the blood pressure pair, see Figure 9, is more linear and elongated than the overall pattern (correlation of 0.953, *p*_*H*_ = 0.01 and *p*_*F*_ = 0.0023). This is partly due to the fact that many points coincide because of discretization (rounding) effects. In the height-weight pair for the same center we see a pattern of negative association (correlation of −0.345, *p*_*H*_ = 0.026 and *p*_*F*_ = 0.0093), possibly driven by only one point.

In center C52, 5 diastolic-systolic observations collapse to 3 perfectly aligned points, resulting in extreme correlations and *p*-values (*p*_*H*_ = 4.910^−13^ and *p*_*F*_ = 5.0610^−13^). This points to a general phenomenon: extreme correlations are sometimes obtained from a combination of small center sizes and overlapping datapoints due to discretisation (in this example: to multiples of 10 mmHg).

In summary, centers can be detected if their correlation is markedly outlying in either the positive or the negative direction with respect to the general tendency across centers and taking into account the center size. In most cases the two tests yield the same conclusion.

In rare cases however, they disagree. One such example is center B102 for the height-weight pair, which is detected with the Fixed margin test (size 146, correlation of 0.4221, *p*_*H*_ = 0.040), but not with the Fisher test (*p*_*F*_ = 0.163).

## 6 Discussion

In this manuscript we have presented two methods for detection of outlying correlations in a multicenter clinical trial. The methods have a different rationale with somewhat different modelling assumptions, but both rely on random effects.

Random effects are essential in both methods and allow to account for heterogeneity in centers above and beyond the variability already seen in the estimates of the Pearson correlation. Without these random effects, this heterogeneity could easily be mistaken for a signal and cause additional false positive detections. In the basketball example, the random effect above and beyond the Fisher variability based on center size is very small. Without this additional component, the p-values in the Fisher scale test become only slightly more significant.

While the modelling assumptions at the basis of the Fisher test, discussed in Section 2, allow for center specific standard deviations in both variables, the underlying modeling assumptions for the Fixed margin test imply that the centers should, at least approximately, have the same scale for this method to be applicable. This is an important point when evaluating both methods. In the simulation setup, standard deviations were kept constant across trials while in real data sets it may not be the case.

It should be stressed that real life data often have a more complex structure than accounted for in our simple models, based on (1), even with center-specific parameters. For example, the correlation coefficients *r*_*X,Y*_ and marginal standard deviations *s*_*X*_ or *s*_*Y*_ might be correlated in their own right (this was actually the case in the baseball players data set, with a significant correlation of 0.4874 between 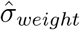 and *r*_*height,weight*_). It is therefore important to realize that the behaviour and performance of these tests depends on the adequacy of the model assumptions to the data at hand, leading, e.g., to different conclusions from the tests in a few cases. In addition, real data may exhibit outliers, discretisation phenomena, deviation from normality, coexistence of different units of measurement across centers etc. These may all point to data quality issues, to be handled manually up front, or to be considered in a broader perspective of CSM.

The applications on fabricated and clinical data sets confronted us with the fact that outlyingness in terms of the correlation, more often than not is accompanied by outlyingness in terms of univariate features of the data, e.g. in the means or standard deviations. This also brings us to the concept of CSM, the process in which the correlation tests are typically embedded. The aim of CSM is to combine different statistical tests addressing different types of inconsistencies to flag problematic centers in an unsupervised fashion. Each test assigns a *p*-value to individual centers, resulting in a huge matrix of *p*-values. These *p*-values in turn provide the basis for the computation of a summarizing score across tests and across multiple variables^1^. In that sense, properties of the *p*-values of individual tests are important. It is also in the scoring stage that tests may be discarded, for instance if a too large percentage of the centers turns out to be flagged, and this property allows to deal with the issue of excessive power at large contamination rates mentioned before.

In summary, we have presented two proposals for a correlation test with demonstrated potential for application in a CSM workflow, as well as in standalone applications. Our simulations indicated largely equivalent performance of both tests.

## Data Availability

We reanalysed data that do not belong to us

## Acknowledgements

To typeset an “Acknowledgements” section.

## Supplemental material

**Figure S1.**
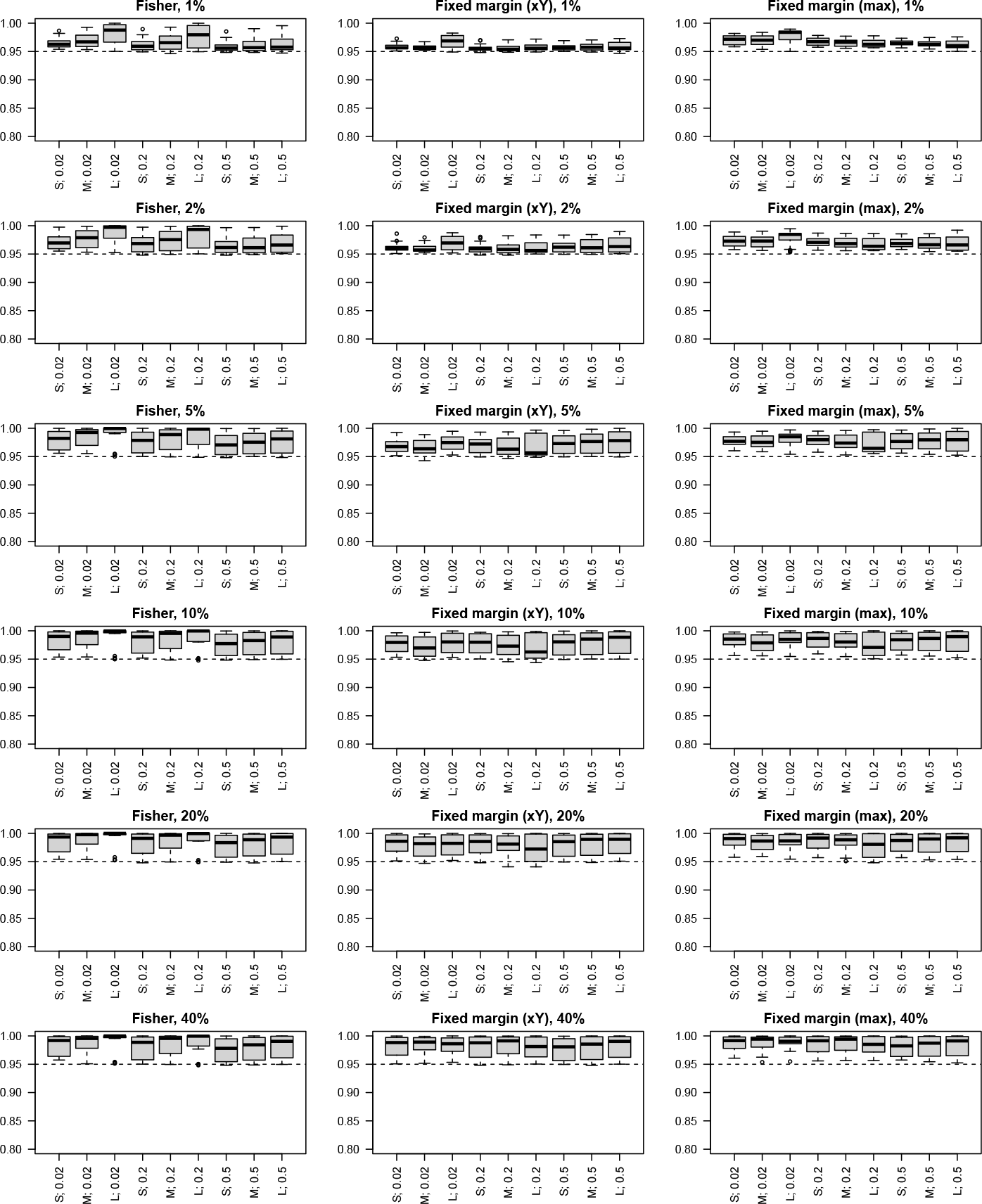
Specificity boxplots in a matrix for different levels of contamination (rows) for Fisher method (1st column) and fixed margin method (2nd column: fixing X; 3rd column: maximum version). Individual boxplots per panel are identified by X-axis labels: center size indicator (Figure 2) followed by Fisher scale standard deviation.

**Figure S2.**
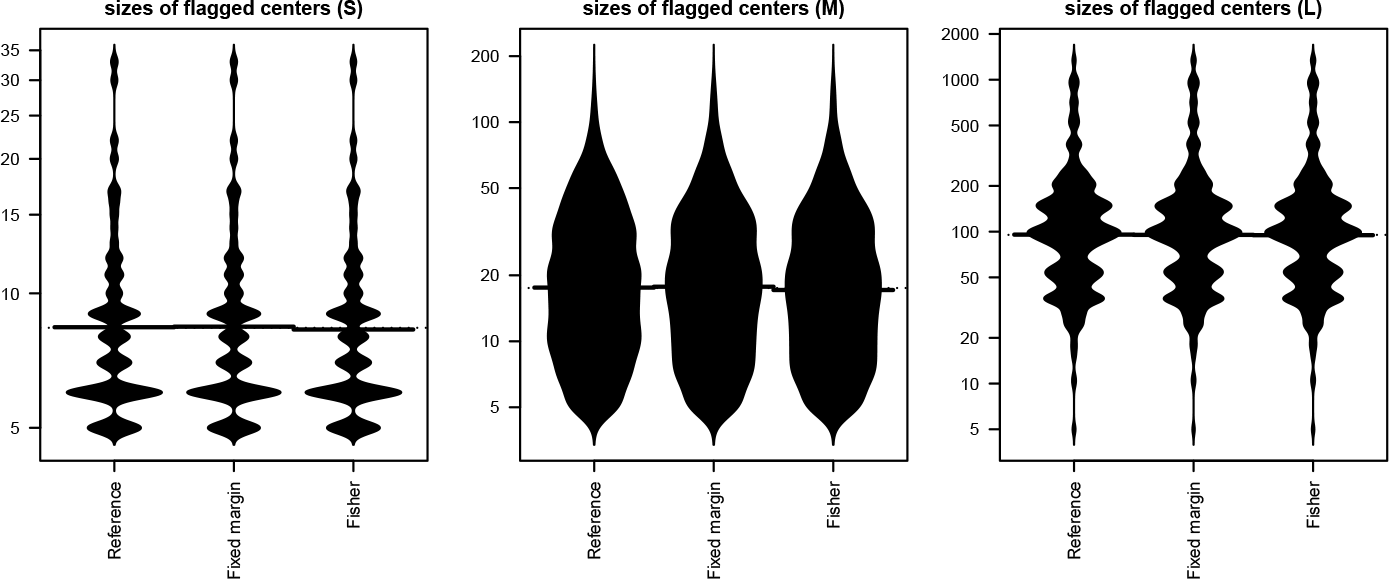
The three panels correspond to the sizes distribution used in the simulation study (left: Small, middle: Medium, right: Large). Within each panel, violin plots compare the sizes of all detected centers (middle: fixed margin method (max), right: Fisher method) with the baseline size distribution (left).

**Figure S3.**
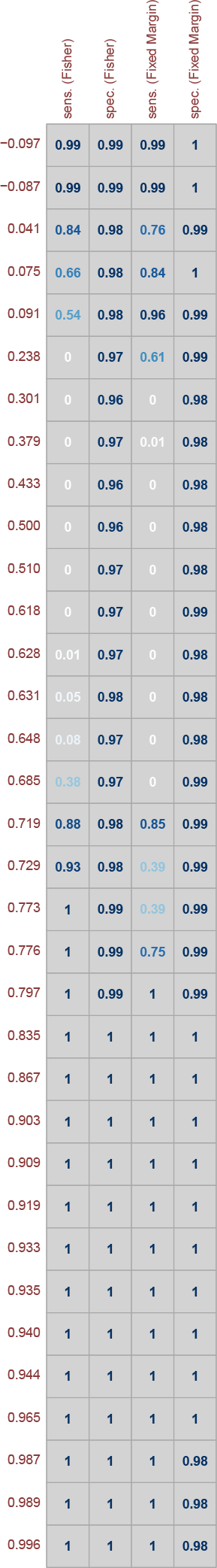
Power and specifity when combining a fabricated dataset with simulated datasets according to the true null model in the first exercise (height-weight). The 34 fabricated scenarios are sorted from smallest correlation to largest correlation (leftmost column).

